## Supplemental Information for "Genetic Nurture in Intergenerational Transmission of Substance Use" (from another version of this preprint: version 1, 2025-08-29)

### Supplementary Information

#### Supplementary Methods

##### Participants

This study included a total of 19,233 genotyped adult offspring who had at least one parent genotyped, comprising 15,966 parent-offspring pairs and 3,267 complete mother-father-offspring trios. For the analysis of overall genetic nurture effects using mixed-effect regression modeling, the sample size varied depending on the availability of substance use data, with a maximum of 15,863 (13,411 parent-offspring pairs and 2,452 complete trios). For parent-of-origin and mediation analyses using structural equation modeling (SEM), full-information maximum-likelihood estimation (FIML) was applied to handle missing data[1]. This approach enabled the inclusion of all 19,233 genotyped adult offspring.

##### Quality control and imputation of genotype data

Lifelines participants were genotyped using three different arrays: the Illumina CytoSNP-12v2 array, the Infinium Global Screening Array® (GSA) MultiEthnic Disease Version 1.0, and the FinnGen Thermo Fisher Axiom® custom array, in order of release. Quality control (QC) of marker and samples were performed separately per array. For the CytoSNP array (released in 2020), quality control (QC) involved filtering SNPs with a minor allele frequency (MAF) above 0.001, a Hardy-Weinberg equilibrium (HWE) p-value >1e-4. A call rate threshold of 0.95 was used for both markers and samples. Sample QC included principal component analysis (PCA) to detect population outliers, and removal of duplicates, individuals with high heterozygosity and ambiguous sex, resulting in 249,249 markers and 15,422 samples. The UMCG Genetics Lifelines Initiative (UGLI) release 1 underwent a two-step QC for marker and sample missingness thresholds (from <80% to <99%), removing monomorphic markers (MAF = 0) and those with HWE p-value >1e-6.

Samples with heterozygosity >4 standard deviations from the mean, duplicates, and ambiguous sex were excluded, yielding 548,029 markers and 36,339 samples. The UGLI release 2 (Affymetrix array) followed a similar two-step QC for call rates (first <80%, then <99%), removing markers with HWE p-value >1e-10 and MAF <0.02, and excluding samples with heterozygosity >4 SD from the mean, duplicates, and those with sex or family discrepancies, resulting in 462,731 markers and 28,249 samples. All arrays were imputed using the Haplotype Reference Consortium (HRC) panel (http://www.haplotype-reference-consortium.org) via the Sanger Imputation Service (http://imputation. sanger.ac.uk). Population stratification was examined using PCA with samples from the 1000 Genomes Project, retaining only individuals of European ancestry. Post-imputation, we filtered for imputation quality (INFO >0.8) and MAF >0.05 and selected high-quality markers (HapMap3+) in each array. We then selected markers that were available in all three arrays, resulting in 1,161,061 common markers. This overlap allowed for reliable haplotype comparison across parent-offspring pairs in the sample of 19,235 offspring with at least one genotyped parent, before matching with phenotype data. Detailed QC reports for each array are available on the Lifelines wiki (http://wiki-lifelines.web.rug.nl/).

**Polygenic scores (PGS) imputation**

Imputation and standardization were performed similarly to the approach described in Kong et al.[2]. For parent-offspring trios and pairs, parental transmitted PGS ($PGS_T$) were computed as the sum of the transmitted maternal and paternal haplotypic PGS. Parental non-transmitted PGS ($PGS_{NT}$) were calculated by summing the scores from both parents. In parent-offspring pairs where data for one parent was missing, the missing $PGS_{NT}$ value for that parent was imputed using the mean score of all available scores from parents of the same type (e.g., the mean paternal PGS was used if the father's data was missing, and the mean maternal PGS if the mother's data was missing) before summing the scores.

#### Statistical analysis

##### *Mediation pathways via parental substance use*

Maternal and paternal mediation pathways were modeled separately due to estimation failure when both maternal and paternal smoking quantity were included in the same models. This failure was attributed to low coverage in pairwise combinations of the predictor, mediator, and outcome variables, despite using Full Information Maximum Likelihood (FIML) to handle missing data.

To evaluate whether the constraints in our mediation models were appropriate, we additionally fitted a saturated model by allowing bidirectional covariances between all variables. This comparison ensures that the hypothesized mediation structure is not overly restrictive while still providing a meaningful and interpretable model of the relationships among variables.

#### Supplementary Results

**Table S1.** Correlations among parental polygenic scores (PGS) and offspring substance use outcomes in Lifelines

| | SmkInit | CigDay | Pack-years | DAlc | CanU | $PGS_{smkInit\_M}$ | $PGS_{smkInit\_P}$ | $PGS_{CigDay\_M}$ | $PGS_{CigDay\_P}$ | $PGS_{DrnkWk\_M}$ | $PGS_{DrnkWk\_P}$ | $PGS_{CanU\_M}$ | $PGS_{CanU\_P}$ |
|---|---|---|---|---|---|---|---|---|---|---|---|---|---|
| SmkInit | 1 | | | | | | | | | | | | |
| CigDay | - | 1 | | | | | | | | | | | |
| Pack-years | - | 0.76*** | 1 | | | | | | | | | | |
| DAlc | 0.21*** | 0.07*** | 0.07*** | 1 | | | | | | | | | |
| CanU | 0.27*** | 0.02 | -0.07*** | 0.19*** | 1 | | | | | | | | |
| $PGS_{smkInit\_M}$ | 0.13*** | 0.04 | 0.02 | 0.01 | 0.04 | 1 | | | | | | | |
| $PGS_{smkInit\_P}$ | 0.11*** | 0.10** | 0.12** | 0.02 | 0.02 | 0.04* | 1 | | | | | | |
| $PGS_{CigDay\_M}$ | 0.02 | 0.11** | 0.10** | -0.01 | 0.03 | 0.13*** | 0.00 | 1 | | | | | |
| $PGS_{CigDay\_P}$ | 0.02 | 0.16*** | 0.10** | 0.01 | -0.02 | 0.03 | 0.10*** | 0.04* | 1 | | | | |
| $PGS_{DrnkWk-M}$ | 0.03 | -0.03 | 0.01 | 0.08*** | 0.03 | 0.25*** | -0.01 | 0.00 | 0.02 | 1 | | | |
| $PGS_{DrnkWk\_P}$ | 0.04 | 0.04 | 0.04 | 0.05* | 0.00 | 0.00 | 0.25*** | -0.06** | 0.05* | 0.03 | 1 | | |
| $PGS_{CanU\_M}$ | 0.05* | -0.05 | -0.03 | 0.01 | 0.05 | 0.20*** | -0.02 | -0.03 | 0.00 | 0.16*** | 0.02 | 1 | |
| $PGS_{CanU\_P}$ | 0.02 | 0.01 | -0.02 | 0.01 | 0.07** | -0.02 | 0.18*** | -0.02 | -0.04* | -0.03 | 0.16*** | 0.02 | 1 |

Note. SmkInit = smoking initiation; CigDay = cigarettes per day; DAlc = daily alcohol intake; CanU = lifetime cannabis use; DrnkWk = drinks per week. CigDay and pack-years are defined only for individuals who have ever smoked, meaning never-smokers have missing data on these variables; therefore, correlations involving them are not available.

Sample sizes for substance use outcomes in offspring ranged from 5,969 to 15,937. Parental PGS was calculated as the sum of transmitted and non-transmitted PGS for each parent, based on the complete trio sample ($N$ = 3,267). Pearson's correlation was used for continuous-continuous variable correlations, point-biserial correlation for binary-continuous correlations, and the phi coefficient for binary-binary correlations. Significance levels: .$*p$ < .05; $**p$ < .01; $***p$ < .001.

**Table S2.** Covariance estimates of maternal and paternal PGS in the parent-of-origin effects analysis

| | **Maternal $PGS_{T\_CigDay}$** | | | **Maternal $PGS_{NT\_CigDay}$** | | |
|---|---|---|---|---|---|---|
| | *β* (SE) | 95% CI | *p* | *β* (SE) | 95% CI | *p* |
| **Paternal $PGS_{T\_CigDay}$** | 0.006 (0.007) | -0.009, 0.019 | 0.421 | 0.012 ( 0.009) | -0.006 , 0.029 | 0.171 |
| **Paternal $PGS_{NT\_CigDay}$** | 0.006 (0.010) | -0.014, .0.025 | 0.570 | -0.005 ( 0.017 ) | -0.040 , 0.026 | 0.753 |

Note. Structural equation modeling was used to test parent-of-origin effects, while accounting for covariances between maternal and paternal transmitted ad non-transmitted polygenic score due to potential genetic assortative mating on smoking. As the analyses used a full information maximum likelihood (FIML) approach to handle missing data, retaining all available data from the full genotyped family sample (*N*=19,233), there was no list-wise *N* for the sample. Standardized coefficients (*β*), bootstrapped standard errors (SE), bootstrapped 95% confidence intervals (CIs) and *p* value were reported.

**Table S3.** Model Fit Comparison: mediation models versus saturated models

| **Model** | **$\chi^2$** | **df** | ***p-value*** | **CFI** | **TLI** | **RMSEA** | **SRMR** | **AIC** | **BIC** |
|---|---|---|---|---|---|---|---|---|---|
| **Maternal cigarettes per day** | | | | | | | | | |
| Mediation model | 0 | | | 1 | 1 | 0 | 0 | 262232.320 | 262444.658 |
| Saturated model | 8.666 | 1 | .003 | .990 | .844 | .020 | .005 | 262238.985 | 262443.459 |
| **Maternal pack-years** | | | | | | | | | |
| Mediation model | 0 | | | 1 | 1 | 0 | 0 | 262877.315 | 263089.654 |
| Saturated model | 8.738 | 1 | .003 | .996 | .941 | .020 | .005 | 262884.054 | 263088.528 |
| **Paternal pack-years** | | | | | | | | | |
| Mediation model | 0 | | | 1 | 1 | 0 | 0 | 248591.166 | 248803.504 |
| Saturated model | 8.722 | 1 | .003 | .996 | .938 | .020 | .005 | 248597.888 | 248802.361 |

Note. $\chi$2 = chi square goodness of fit statistic; df = degrees of freedom; CFI = Comparative Fit Index; TLI = Tucker Lewis Index; RMSEA (90% CI) = Root-Mean-Square Error of Approximation with 90% confidence intervals; SRMR = Standardized Square Root Mean Residual; AIC =Akaike information criterion; BIC = Bayesian Information Criterion.

**Table S4**: Summary statistics of Genome-Wide Association Studies used to calculate polygenic scores

| **Phenotype** | **Abbrev.** | $N_{Total}$ | **Number of independent SNPs** | **SNP heritability (SE)** | **Citation** |
|---|---|---|---|---|---|
| Smoking Initiation | SmkInit | 2,669,029 | 1,752 | 0.08 (0.002) | Saunders et al. 2023 |
| Cigarettes Per Day | CigDay | 618,489 | 153 | 0.08 (0.005) | Saunders et al. 2023 |
| Drinks Per Week | DrnkWk | 2,428,851 | 501 | 0.04 (0.001) | Saunders et al. 2023 |
| Lifetime Cannabis Use | CanU | 184,765 | 8 | 0.11 (0.01) | Pasman et al. 2018 |
